# RACIAL AND ETHNIC DIFFERENCES IN LONG-TERM CARDIOVASCULAR MORTALITY AMONG WOMEN AND MEN FROM THE CAC CONSORTIUM

**DOI:** 10.1101/2024.03.01.24303634

**Authors:** Shmuel Rosenblatt, Michael J. Blaha, Ron Blankstein, Yvette Yeboah-Kordieh, Khurram Nasir, Fay Lin, Daniel S. Berman, Michael D. Miedema, Seamus P. Whelton, John Rumberger, Matthew J. Budoff, Jonathon Leipsic, Leslee J. Shaw

**Author notes:** Address for correspondence: Leslee Shaw, 1425 Madison Avenue, Room L2-33, Icahn School of Medicine at Mount Sinai, New York, New York, 10029.

## Abstract

**Background:** Despite an increasingly diverse population, knowledge regarding racial and ethnic disparities is limited among women and men undergoing atherosclerotic cardiovascular (ASCVD) screening. Our aim was to compare CV mortality by ASCVD risk and coronary artery calcium (CAC) scores among Black and Hispanic women and men compared to other participants.

**Design and Methods:** From the CAC Consortium, 42,964 participants with self-reported race and ethnicity were followed for a median of 11.7 years. Multivariable Cox proportional hazards regression models were used to estimate CV mortality, with separate analyses by sex.

**Results:** One-third of enrollees were women; 977 self-reported as Black, 1,349 as Hispanic, 1,621 as Asian, and 740 as American Indian/Native Alaskan/Hawaiian or other; the remainder were white. Black women and men had higher ASCVD risk and CAC scores yielding the highest CV mortality compared to other participants. Among Black women and men with a 0 CAC or ASCVD risk score <5%, hazard ratios (HRs) were 6-9-fold higher than that of other women and men. In men with CAC scores ≥100, Black men (HR: 4.2,p<0.001) had the highest CV mortality compared to all other men. A similar high-risk pattern was noted for Black women with CAC scores ≥100 (p<0.001), even when adjusting for the ASCVD risk score. Overall, Hispanics had an elevated CV mortality, higher than others but less than that of Black participants. Patterns of intermediate risk were notable for Hispanic men with a CAC score of 0 (HR=3.6, p=0.006) and ≥100 (HR=2.3, p=0.03).

**Conclusions:** The disproportionately high and excess CV mortality among Black women and men represents significant barriers to reducing the burden of ASCVD through effective screening using ASCVD risk and CAC scores.

---

Computed tomographic imaging of coronary artery calcium (CAC) is commonly used in screening of asymptomatic individuals at-risk for atherosclerotic cardiovascular disease (ASCVD).^1–3^ There are numerous multicenter and population registries reporting a graded relationship between the CAC score and CV outcomes, yet often these reports lack racial and ethnic diversity.^4–7^ Available evidence is often divergent as to whether individuals of diverse race and ethnicity have a higher or lower burden of CAC.^8,9^ Moreover, unique sex differences have been reported whereby women have a lower burden of CAC but with more extensive CAC, including those with multivessel involvement, women are at higher risk of CV mortality.^3^ Thus far, reports have focused largely on the risk gradient with increasing CAC scores among Black and Hispanic participants, without comparative analyses of the influence of sex on race and ethnicity on CV outcomes.^1,8,9^

The CAC Consortium is a large multicenter registry focusing on long-term mortality associated with CV screening using the atherosclerotic CV risk and CAC score.^2^ In the current analysis, we examined unique sex differences in long-term CV mortality by CAC scores and risk factors among diverse race and ethnic subgroups of the CAC Consortium.

## Methods

### CAC Consortium Enrollment and Study Population

Details of the CAC Consortium have been previously published^2,3^ however, in brief, a total of 66,636 patients were enrolled from 4 centers from 1993-2014. Participants were included in this study if they were ≥18 years of age, asymptomatic, and had no history of CAD at the time of their index CAC scan. For this report, we included 42,964 participants (from 3 of the 4 sites) with self-reported race and ethnicity. From this subset, 977 self-reported as Black, 1,349 as Hispanic, 1,621 as Asian, 740 as other (i.e., 165 self-reported as American Indian, Alaskan Native, or Native Hawaiian / Pacific Islander, and 581 self-reported as other), and 38,277 reported as white. Previous reports serve as the foundation for the current analysis.^3,9^

### Data Collection for CAD Risk Factors and the ASCVD Risk Score

Traditional CAD risk factors and laboratory tests, when available, were obtained at the time of CAC scans. Hypertension was defined as a previous diagnosis or treatment with antihypertensive medication. Dyslipidemia was defined as either a previous diagnosis of hyperlipidemia or treatment with lipid lowering medications. Diabetes was defined as a previous diagnosis or treatment with hypoglycemic medications or insulin. Treatment was differential for each risk factor and was aggregated into hypertensive, diabetic, and dyslipidemia medications. Family history was defined as having at least one first-degree relative with CVD, with exception, the center in Columbus, which defined it as having a family member with a premature history of CVD (<55 years of age for men, <65 years of age for women). The 10-year ASCVD risk score was also calculated, as described in the design manuscript.^2^

### Computed Tomography CAC Scan Protocol and Interpretation

CT scans were obtained using non-contrast cardiac gated CT scans at each individual center using standardized protocols.^2^ A CAC scan was interpreted using the Agatston scoring method.^10^ A CAC scan was categorized with scores of 0, 1-99, and ≥100. Analysis focused on subgroups with a 0 score and for those with a CAC score ≥100. Few participants had CAC scores of 400 or higher and to allow for examination of sex, race, and ethnic differences, we selected a threshold a CAC score ≥100.

### Death Ascertainment and Categorization

Details of the methodology for ascertainment of death and causality are previously reported.^2^ Ascertainment of death was done using the Social Security Death Index Death Master File. Follow-up and ascertainment of death was completed through June 1^st^, 2014. Cause of death was found in the National Death Index. ICD-9 or ICD-10 codes were used to report the cause of death including CV, cancer, pulmonary, and injury, to name a few. The primary endpoint for this report was CV mortality. The median time to CVD death was 11.7 years (interquartile range: 9.9-13.1 years).

### Statistical Methods

Women and men were largely examined in separate analyses due to their variable frequency of CAC and CV mortality. Descriptive statistics were compared across the diverse race and ethnic subgroups of women and men using a chi-square for frequency and nonparametric tests for continuous data. Age is presented as median with interquartile range values. The focus of this report was to highlight differences among Black and Hispanic women and men, as such, the results are presented with these subgroups being the focus of all comparisons. For all Cox models, patients in the Asian, American Indian, Alaska Native, Native Hawaiian, or other, and white subgroups had similar CV mortality and are grouped as the comparator. The primary outcome for this analysis was CV mortality. An initial Cox proportional hazards model examined CV survival across race and ethnicity. A first order test for interaction of sex by race / ethnicity was also examined. Final Cox models were examined among participants with ASCVD risk scores >7.5% and with CAC scores ≥100. For all Cox models, the proportional hazards assumption was met. The area under the curve (AUC) from receiving operating characteristic models for classifying CV mortality using the method of DeLong et al.^11^ We compared the 10-year CV death rates across separate racial and ethnic subgroups of women and men with ASCVD scores <5% and 0 CAC scores using Kaplan-Meier survival analysis. For CAC models, all results were similar to the presented findings if the ASCVD risk score, enrolling site, and the year of enrollment was added to the model as covariates.

### Covariates for Referral Bias and Missingness in Race and Ethnicity

We explored potential bias in referral and missingness by examining enrollment patterns among racial and ethnic subgroups. From the 3 sites reporting race and ethnicity, there were 42,964 participants with self-reported race and ethnicity data. From the 3 sites, ∼1 in 4 enrollees had missing race and ethnicity data. Documentation of race and ethnicity varied by year of enrollment with completion rates improving over time. When comparing CV mortality using a dichotomous variable of available versus missing race and ethnicity, the p value was p=0.48 when the ASCVD risk and CAC scores were added to the model as covariates. In our analyses, to correct for selection bias and missingness in race and ethnicity, we included the year of scanning and enrolling site as covariates in all Cox models but also included the ASCVD risk score in prognostic models including the CAC score.

## Results

### Baseline Cardiac Risk Factors and ASCVD Risk Scores

**Table 1** provides descriptive statistics for women and men in the CAC Consortium. Approximately one-third of enrollees were women with a mean age of 54.7 years. Across the diverse groups, women were generally older than men at the time of their CAC scan. It is notable that hypertension was most prevalent in Black women (60.9%) as compared to other subsets of women. Moreover, Black women had a high rate of smoking (13.8%), diabetes (16.1%) and were more often obese (39.8%). Hispanic women also had a high rate of diabetes (18.3%), with few Black and Hispanic women having documented treatment for diabetes. A similar pattern was noted among men where half of Black participants had hypertension (50.9%), a rate higher than for other male subgroups. Additionally, diabetes was more common among Black (18.1%) and Hispanic (18.1%) men, higher than for other subgroups. Overall, Black participants were the least likely to have a recorded family history of premature CVD among the subgroups in this cohort (49.6% for women and 37.5% for men).

**Table 1.** Descriptive Statistics for Traditional CV Risk Factors in Women (n=15,023) and Men (n=27,941) from the CAC Consortium. Data are presented as %, except for age which is reported with a median (interquartile range=25^th^, 75^th^ %ile).

| **Women**<br>(n=15,023) | **Black**<br>(n=391) | **Hispanic**<br>(n=520) | **Other**<br>(n=235) | **Asian**<br>(n=589) | **white**<br>(n=13,288) | p value |
| --- | --- | --- | --- | --- | --- | --- |
| Age (in years) | 55 (48, 64) | 54 (46, 61) | 55 (49, 62) | 54 (48, 62) | 56 (50, 63) | <0.001 |
| Hypertension | 60.9% | 36.9% | 30.2% | 37.4% | 29.9% | <0.001 |
| Smoking | 13.8% | 6.5% | 15.7% | 5.3% | 8.9% | <0.001 |
| Diabetes Mellitus | 16.1% | 18.3% | 2.6% | 11.7% | 4.9% | <0.001 |
| Dyslipidemia | 56.8% | 55.6% | 54.0% | 57.9% | 62.5% | <0.001 |
| Family History of Premature CVD | 49.6% | 51.3% | 61.3% | 51.8% | 56.6% | <0.001 |
| BMI $\geq 30$ kg/m<sup>2</sup><br>(n=11,647) | 39.8% | 29.1% | 26.8% | 10.5% | 23.2% | <0.001 |
| Hypertensive Treatment<br>(n=3,421) | 42.7% | 34.3% | 0.0%* | 26.4% | 20.8% | <0.001 |
| Aspirin (n=9,938) | 49.2% | 35.5% | 41.6% | 17.1% | 39.3% | <0.001 |
| Dyslipidemia Treatment<br>(n=10,273) | 22.2% | 29.8% | 20.8% | 16.5% | 19.3% | 0.043 |
| Diabetic Treatment<br>(n=287) | 0.0% | 6.7% | 15.0% | - | 4.2% | 0.21 |
| **Men**<br>(n=27,941) | **Black**<br>(n=586) | **Hispanic**<br>(n=829) | **Other**<br>(n=505) | **Asian**<br>(n=1,032) | **white**<br>(n=24,989) | p value |
| Age (in years) | 54 (47, 61) | 52 (45, 59) | 52 (46, 59) | 52 (45, 61) | 53 (46, 61) | <0.001 |
| Hypertension | 50.9% | 38.7% | 24.6% | 37.0% | 28.0% | <0.001 |
| Smoking | 15.7% | 12.2% | 13.5% | 8.8% | 9.9% | <0.001 |
| Diabetes Mellitus | 18.1% | 18.1% | 9.3% | 11.7% | 5.7% | <0.001 |
| Dyslipidemia | 53.4% | 60.3% | 57.6% | 63.1% | 61.3% | <0.001 |
| Family History of Premature CVD | 37.5% | 41.0% | 45.9% | 42.0% | 46.9% | <0.001 |
| BMI $\geq 30$ kg/m<sup>2</sup><br>(n=11,647) | 31.7% | 31.8% | 35.9% | 10.6% | 26.9% | <0.001 |
| Hypertensive Treatment<br>(n=6,383) | 37.4% | 28.7% | 22.2%* | 24.4% | 22.3% | <0.001 |
| Aspirin (n=18,346) | 45.6% | 42.7% | 42.7% | 33.2% | 46.3% | <0.001 |
| Dyslipidemia Treatment<br>(n=18,905) | 23.6% | 32.3% | 23.4% | 27.0% | 25.7% | 0.16 |
| Diabetic Treatment<br>(n=415) | 9.1%* | 25.0% | 0.0%* | 0.0%* | 5.8% | 0.91 |

The p value for age is a nonparametric Kruskal-Wallis statistic. Other includes self-reported Alaskan Indian, Alaska Native, Native Hawaiian / Pacific Islander, or self-reported as "other". \*12 or fewer individuals were reported in this cell.

Across the enrolling years, Black participants were more often referred to CV screening with an ASCVD risk score >7.5% (**Figure 1**). By the ASCVD risk score (**Table 2**), more than 40% of Black women had a score >7.5%, nearly twice as high as for other female subgroups. A similar pattern was noted for Black men where 57.3% had an ASCVD risk score >7.5%. Across all subgroups, ASCVD risk scores were high among Black women and men (average values of 10.4% and 10.9% for Black women and men). Among participants with an ASCVD risk score >7.5%, Black and Hispanic women and men were younger than other enrollees (p<0.001). A subsample of women and men had available risk factor measurements that are presented in **Supplemental Table 1**.

**Figure 1.**
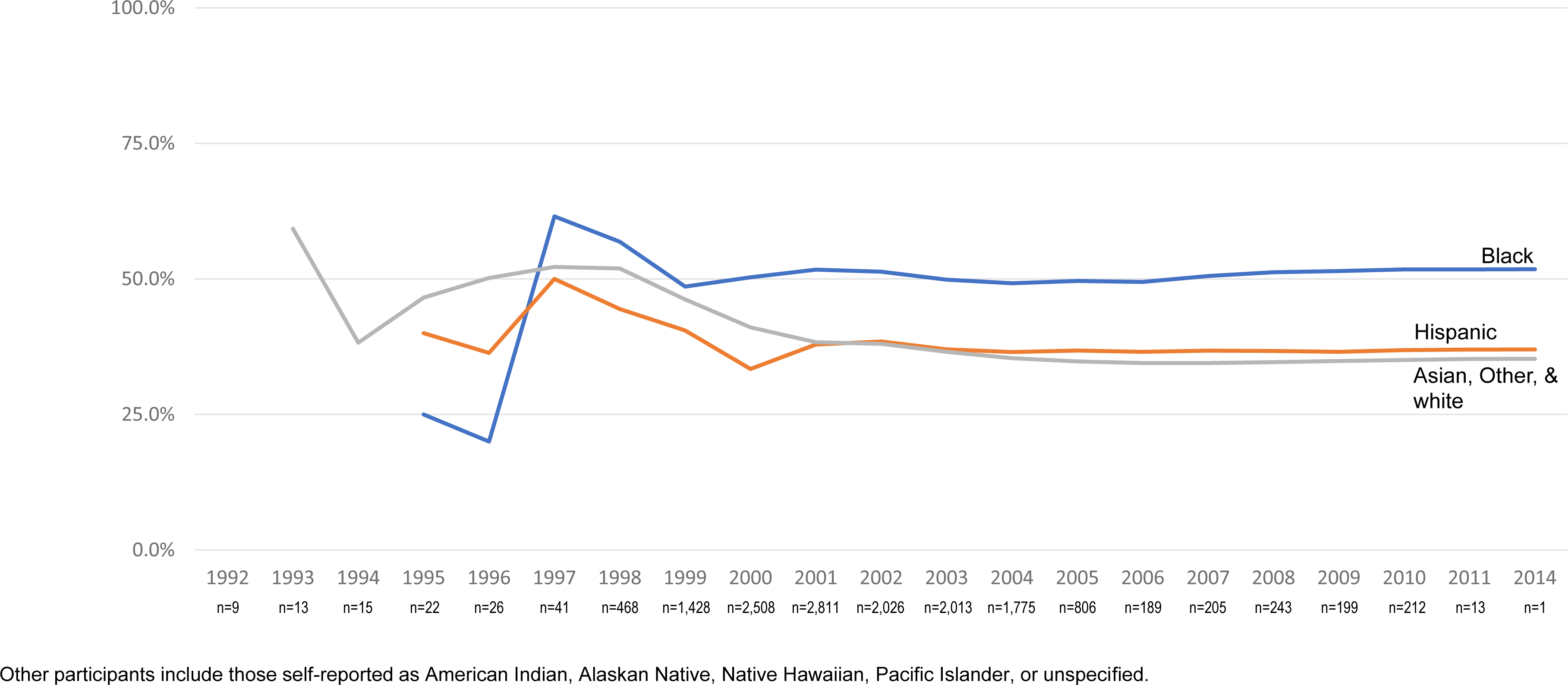
Cumulative Proportion of Participants with an ASCVD Risk Score >7.5% among Black, Hispanic, and Asian, American Indian/Alaskan Native, Native Hawaiian, other, and white participants Across Enrolling Years. The numbers below the x-axis include the number of enrolled participants in that year.

**Table 2.**
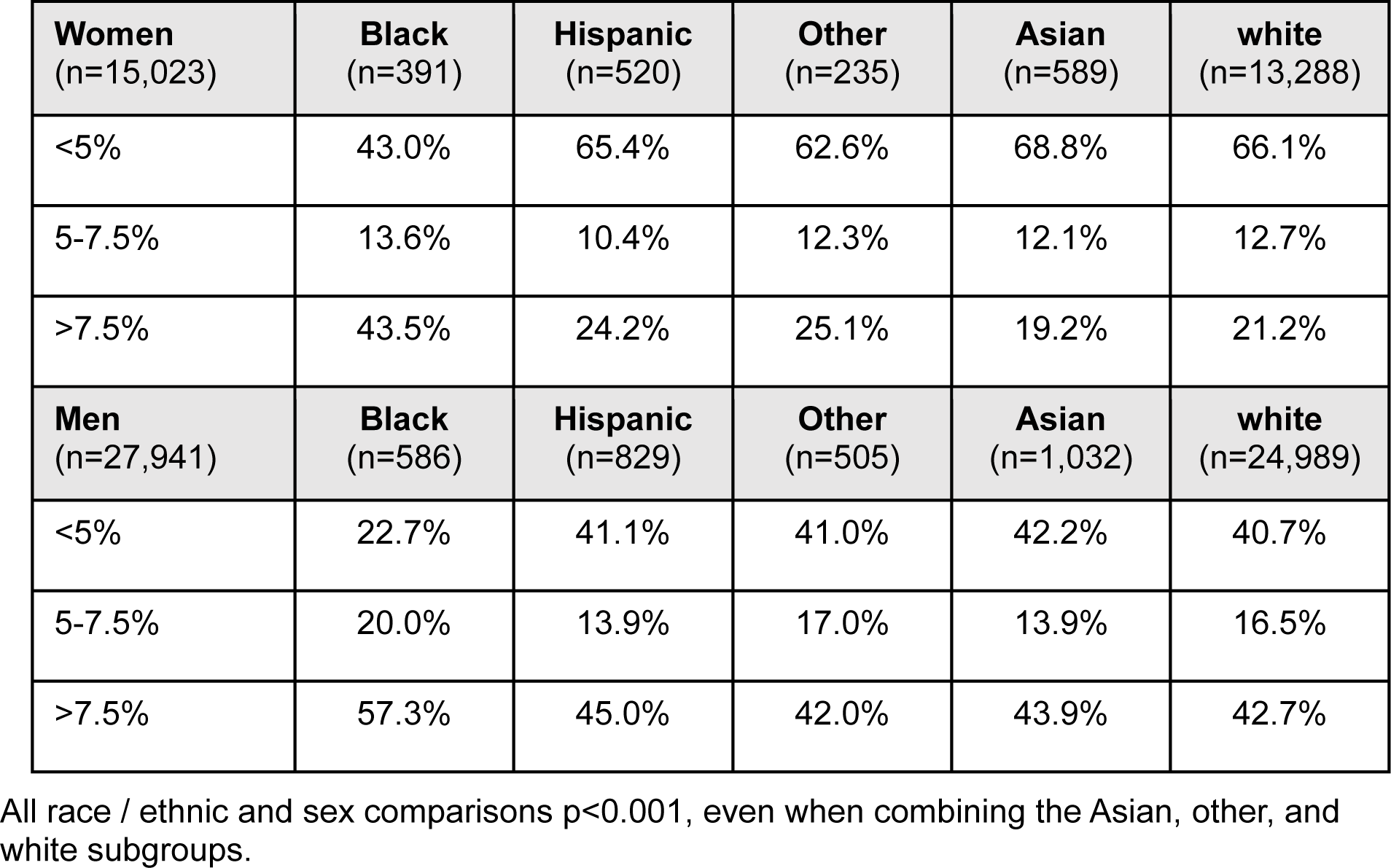
From the CAC Consortium (n=42,964), Proportion of Low to High ASCVD Risk Individuals Among Women and Men of Diverse Race and Ethnicity.

| **Women**<br>(n=15,023) | **Black**<br>(n=391) | **Hispanic**<br>(n=520) | **Other**<br>(n=235) | **Asian**<br>(n=589) | **white**<br>(n=13,288) |
| --- | --- | --- | --- | --- | --- |
| <5% | 43.0% | 65.4% | 62.6% | 68.8% | 66.1% |
| 5-7.5% | 13.6% | 10.4% | 12.3% | 12.1% | 12.7% |
| >7.5% | 43.5% | 24.2% | 25.1% | 19.2% | 21.2% |
| **Men**<br>(n=27,941) | **Black**<br>(n=586) | **Hispanic**<br>(n=829) | **Other**<br>(n=505) | **Asian**<br>(n=1,032) | **white**<br>(n=24,989) |
| <5% | 22.7% | 41.1% | 41.0% | 42.2% | 40.7% |
| 5-7.5% | 20.0% | 13.9% | 17.0% | 13.9% | 16.5% |
| >7.5% | 57.3% | 45.0% | 42.0% | 43.9% | 42.7% |

All race / ethnic and sex comparisons $p < 0.001$ , even when combining the Asian, other, and white subgroups.

### Prevalence of Baseline CAC Scores

Among women, more than half had no detectable CAC, except for Black women where 49.4% had a 0 score (**Table 3**, p<0.001). Nearly two-thirds of Black and Hispanic women with a 0 CAC score were age <55 years, a rate higher than that of other women (p<0.001). Of note, Black women had more prevalent CAC with an odds ratio of 1.52 (95% CI: 1.24-1.86), p<0.001) including 19% with a CAC score of 100 or higher. Among men, across the diverse groups, ∼34%-40% had no detectable CAC (p=0.12). Nearly one-third of Black men had CAC scores of 100 or higher.

**Table 3.** Frequency of CAC Score Subsets Among Women and Men of Diverse Race and Ethnicity Enrolled in the CAC Consortium (n=42,964). Using a logistic regression model, Black women had an odds ratio (OR) of 1.52 (95% CI: 1.24-1.86, p<0.001) for CAC >0 as compared to other racial and ethnic subsets of women.

| **Women (n=15,023)** | **CAC 0** | **CAC 1-99** | **CAC ≥100** |
| --- | --- | --- | --- |
| Black (n=391) | 49.4% | 31.7% | 18.9% |
| Hispanic (n=520) | 60.8% | 24.8% | 14.4% |
| Asian (n=589) | 64.5% | 21.2% | 14.3% |
| Other (n=235) | 60.0% | 23.8% | 16.2% |
| white (n=13,288) | 59.7% | 25.8% | 14.5% |
| **Men (n=27,941)** | **CAC 0** | **CAC 1-99** | **CAC 100-399** |
| Black (n=586) | 40.3% | 33.6% | 26.1% |
| Hispanic (n=829) | 36.7% | 36.3% | 27.0% |
| Asian (n=1,032) | 40.1% | 32.8% | 27.1% |
| Other (n=505) | 33.7% | 38.4% | 27.9% |
| white (n=24,989) | 36.0% | 33.3% | 30.6% |

\*p<0.001 for women and p=0.006 for men. When combining Asian, other, and white, the p value is <0.001 for women and 0.04 for men.

### Observed CV Mortality Among Racial and Ethnic Subgroups of Women and Men

Of the 42,964 enrollees, Black and Hispanic participants had an elevated hazard for CV mortality (**Figure 2**). Moreover, there was a significant sex by race / ethnicity interaction with regards to prediction of CV mortality (p<0.001). In separate models by sex, the relative hazard for CVD death was 4.59 (95% CI: 2.78-7.59) for Black women and 2.26 (95% CI: 1.83-3.99) for Hispanic women, as compared to the remaining women. Similarly, the hazard for CVD mortality was 3.75 (95% CI: 2.57-5.47) for Black men and 1.77 (95% CI: 1.14-2.75) for Hispanic men, as compared to the remaining men. Sex, race, and ethnicity were similarly predictive when the enrolling site, year of enrollment, ASCVD risk score, and CAC findings were added to the Cox models as covariates.

**Figure 2.**
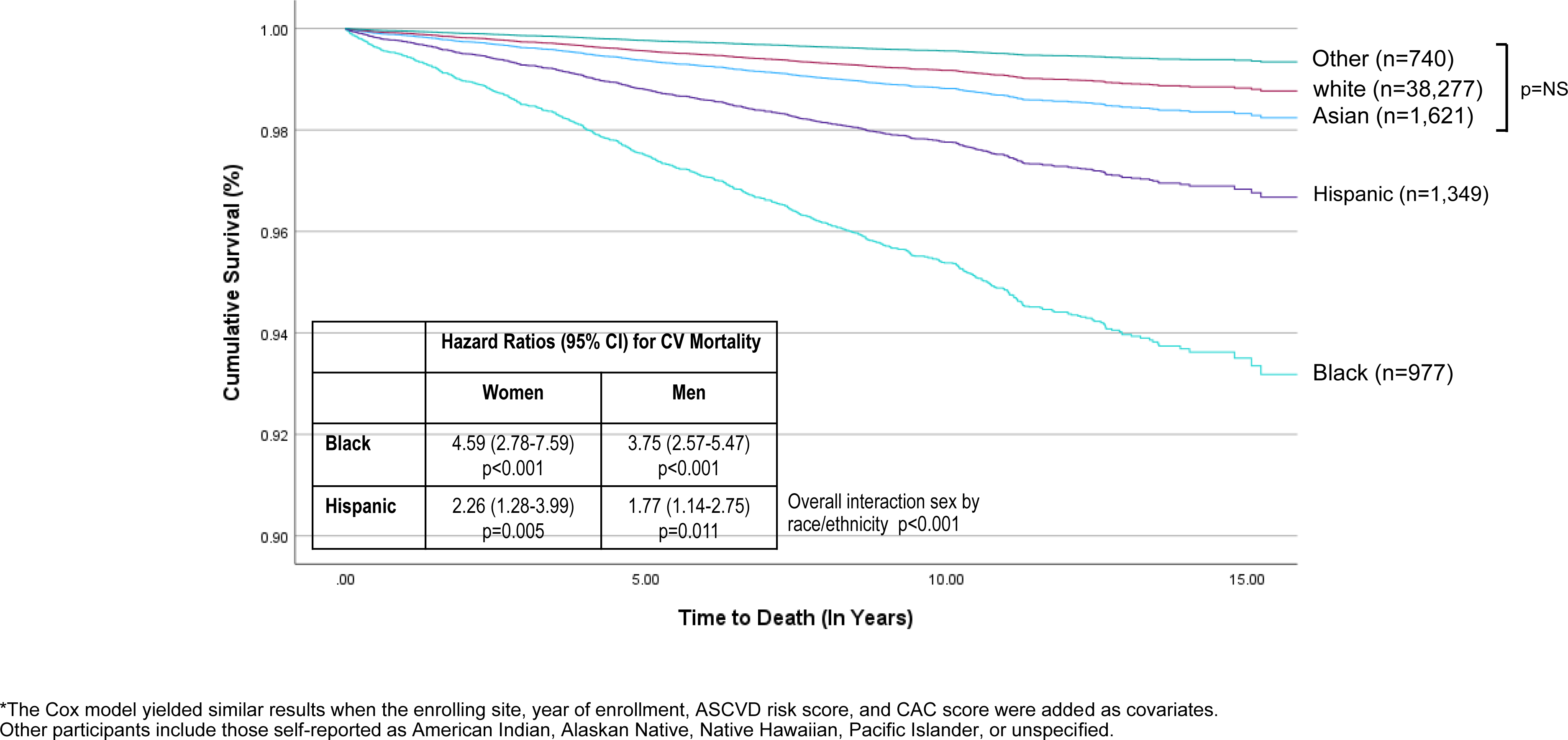
CVD Survival (N=612 deaths) Among Women and Men of Diverse Race and Ethnicity Enrolled in the CAC Consortium (n=42,964). The tables below include the hazard ratios (HR) and 95% confidence intervals (CI) for the CV mortality model reflecting the results from an interaction of sex and race / ethnicity.* CV survival was similar among Asian, white, and other participants.

### CV Mortality Among ASCVD Risk Scores >7.5% and CAC Scores ≥100 by Sex, Race, and Ethnicity

Comparing participants with an ASCVD risk score >7.5%, differences were noted in separate models for women and men across race and ethnicity (**Figure 3**), even when including the enrolling site and year of enrollment as covariates. In **Figure 3**, Black and Hispanic women and men with an ASCVD risk score >7.5% had significantly worse CV mortality as compared to other participants. Of note, despite separate scores assigned to African American race for the ASCVD risk score, Black and Hispanic women with ASCVD risk scores >7.5% had similar CV survival. Across all individuals with ASCVD risk scores >7.5%, women had reduced CV survival (p<0.001).

**Figure 3.**
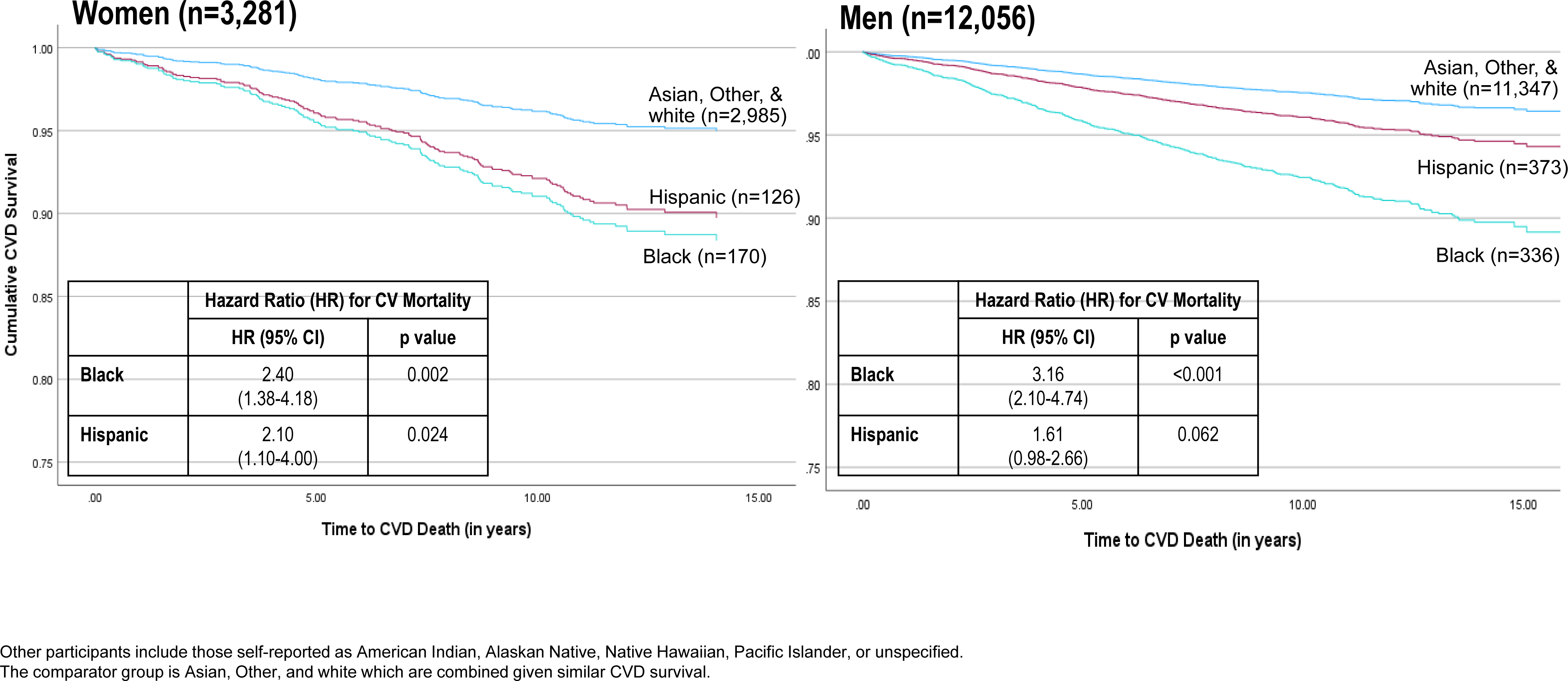
CVD Survival Among Women and Men of Diverse Race and Ethnicity with ASCVD Risk Scores >7.5% (n=15,337). The Cox survival curves are based on separate models for women and men. The tables below include the hazard ratios (HR) and 95% confidence intervals (CI) for CV mortality from models in women and men.

Across all consortia subgroups, a CAC score ≥100 was highly predictive of CV death (p<0.007). Among those with a CAC score of ≥100 (**Figure 4**), there was a similar worsening of CVD mortality among Black and Hispanic women and men as compared to the combined group of other participants. Among women with a CAC score ≥100, the relative hazard for CV mortality was 4.56 (95% CI: 2.50-8.33) and 2.34 (95% CI: 1.08-5.04) for Black and Hispanic women.

**Figure 4.**
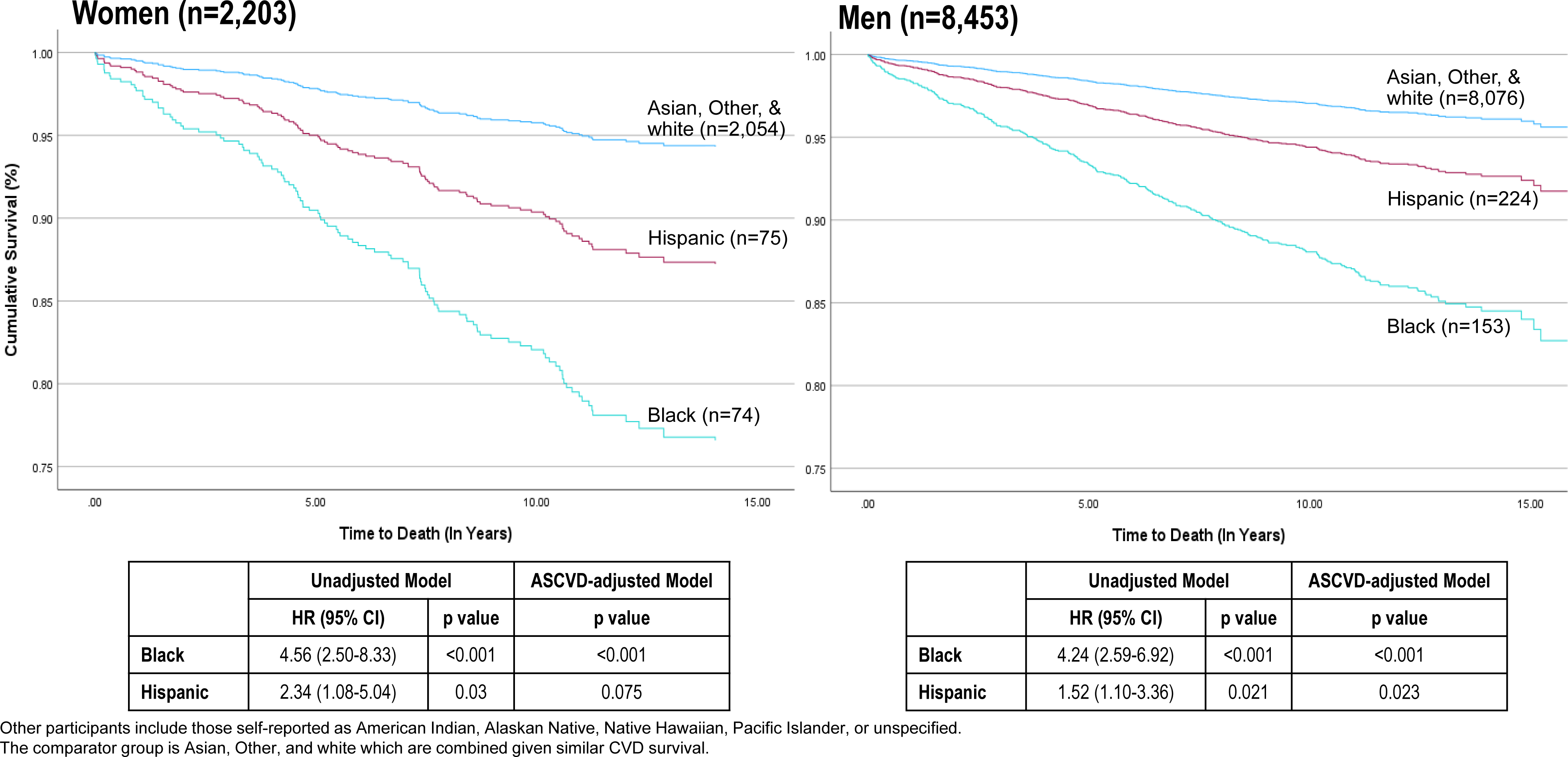
CVD Survival Among Women and Men of Diverse Race and Ethnicity with CAC Scores ≥100 (n=10,656). The Cox survival curves are based on separate models for women and men. The tables below include the hazard ratios (HR) and 95% confidence intervals (CI) for CV mortality from separate models for women and men and included unadjusted and ASCVD risk score-adjusted findings.

Among men with a CAC score ≥100, the relative hazard for CV mortality was 4.24 (95% CI: 2.59-6.92) and 1.92 (95% CI: 1.10-3.36) for Black and Hispanic men. In a model including the ASCVD risk score as a covariate, Hispanic women and men with a CAC score ≥100 no longer had a significantly elevated risk for CV mortality (p=0.074 and p=0.867). Across all individuals with CAC scores ≥100, women had reduced CV survival (p<0.001).

### 10-Year Kaplan-Meier CV Mortality Rates in Low Risk ASCVD Scores <5% and 0 CAC Scores

We also examined differential 10-year CV mortality rates among low-risk participants including those with an ASCVD risk score <5% (**Figure 5a**) and a CAC score of 0 (**Figure 5b**). In a subset of participants with ASCVD risk scores <5% (n=21,140), the 10-year Kaplan-Meier CV mortality rates were similar among women (p=0.22) but varied among men (p<0.001) of diverse race and ethnicity. Black and Hispanic men had 10-year CV mortality of 2.4% (p<0.0001) and 1.5% (p=0.006) when compared to the other male participants. Among low ASCVD risk Black and Hispanic men, hypertension was significantly associated with CV mortality (p=0.002).

**Figure 5.**
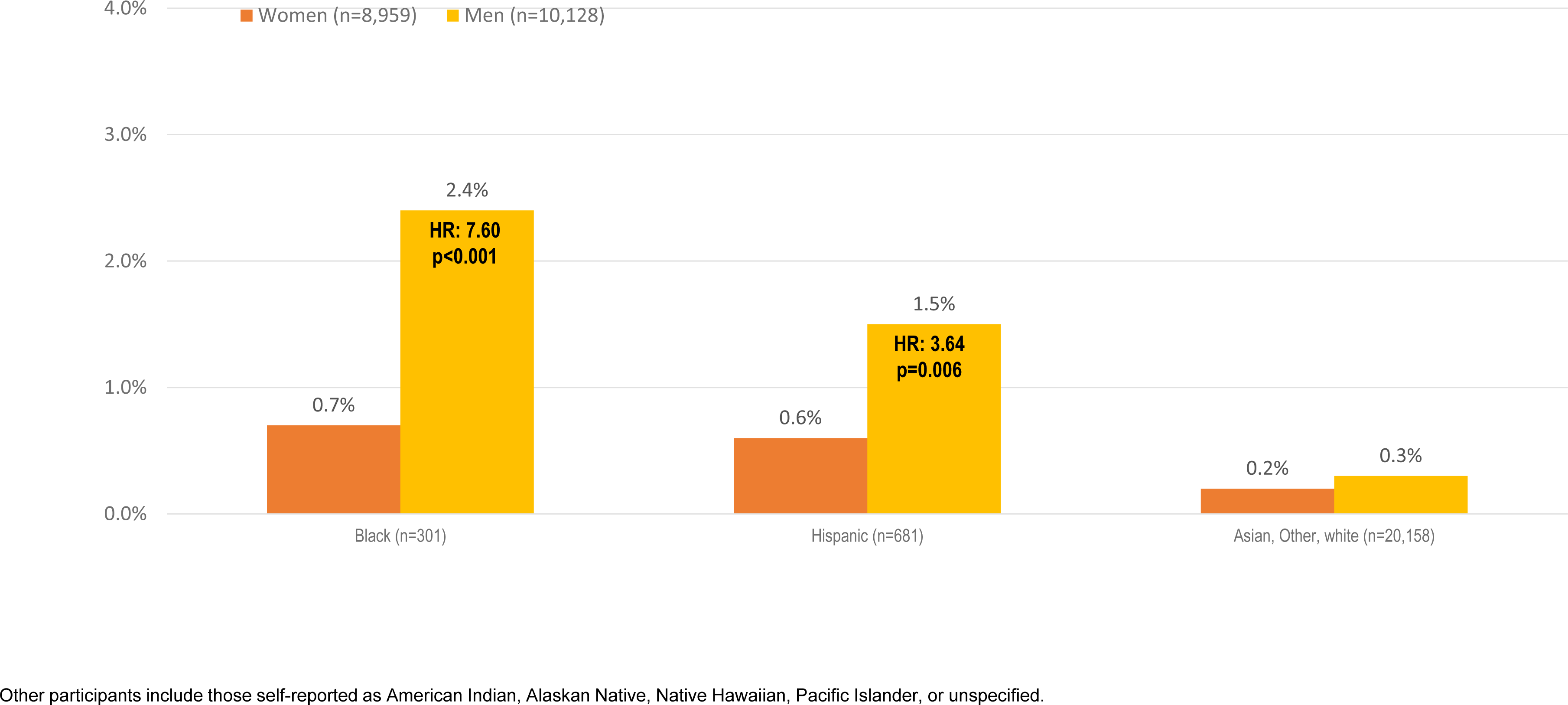
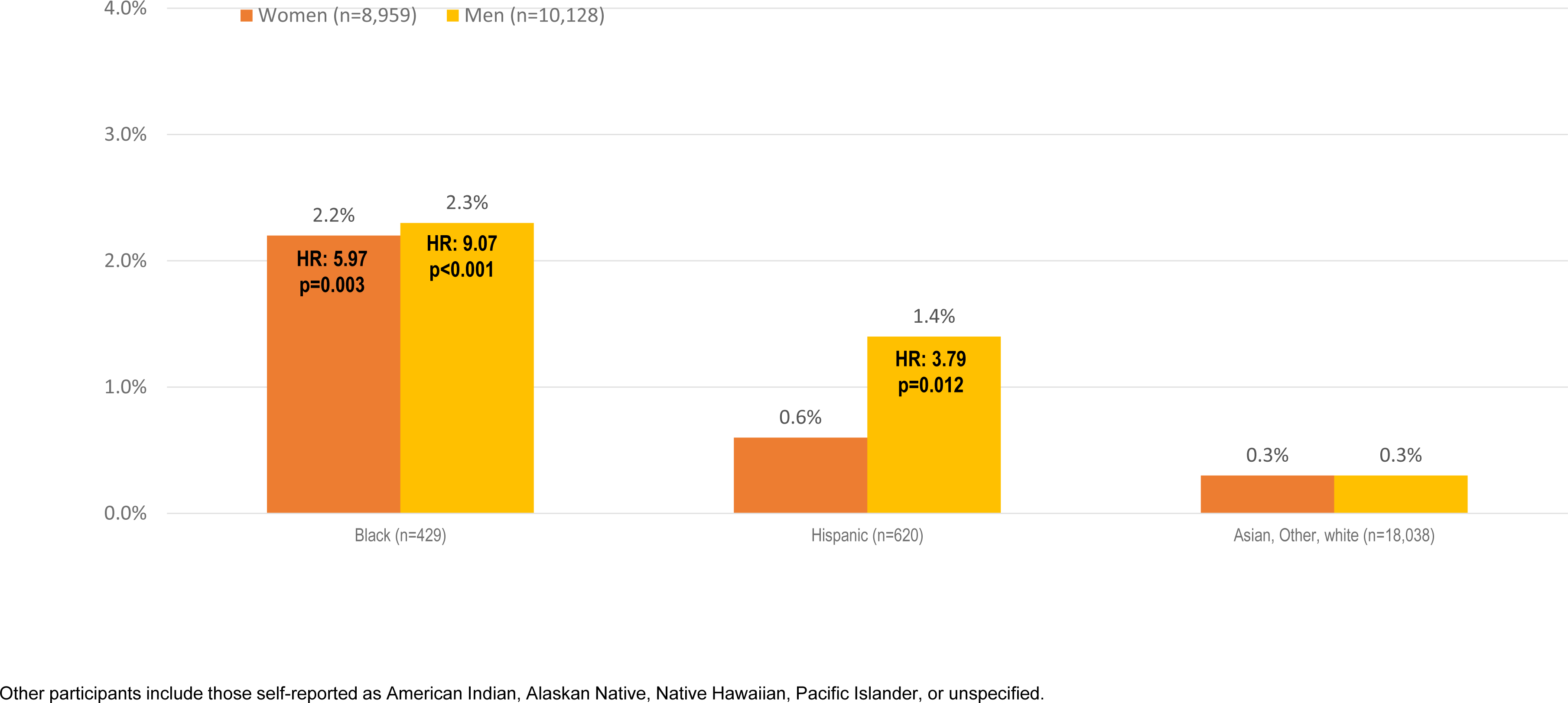
a. 10-year Kaplan-Meier CV Mortality Rates* Among Women and Men of Diverse Race and Ethnicity with an ASCVD Risk Score <5% (n=21,140). The CVD death rates are similar among women (p=0.202) but statistically different among men (p<0.001) of diverse race and ethnicity. The hazard ratios (HR) for CVD mortality in Black and Hispanic men was 7.6 (p<0.001) and 3.6 (p=0.006), as reported at the top of their columns. CV mortality risk remained elevated among Black (p<0.001) and Hispanic (p=0.005) men when the enrolling site and year of enrollment were added to the Cox model as covariates. b. 10-year Kaplan-Meier CV Mortality Rates* Among Women and Men of Diverse Race and Ethnicity with a CAC Score of 0 (n=19,087). Within a Cox model for women, Black women with a 0 CAC score had a hazard ratio (HR) of 6.0 (p=0.003), as reported at the top of their column. Within a Cox model for men, Black and Hispanic men with a 0 CAC score had a HR of 9.1 (p<0.001) and 3.8 (p=0.012), reported at the top of the column. CV mortality risk remained elevated among Black women (p=0.034) and Black (p<0.001) and Hispanic (p=0.014) men when the ASCVD risk score, enrolling site, and year of enrollment were added to the Cox model as covariates.

Among participants with CAC scores of 0 (n=19,087), Black women and Black and Hispanic men had significantly elevated 10-year CVD mortality. Among Black women with a 0 CAC score, the hazard for CVD mortality was 5.9 (p=0.003). Similarly, Black and Hispanic men with a 0 CAC score had a higher 10-year CV mortality of 2.3% (p<0.001) and 1.4% (p=0.004) when compared to other male participants. Among Black and Hispanic men with a 0 CAC score, age ≥65 years was significantly associated with CV mortality (p=0.005). Among Black women with a 0 CAC score, diabetes (p=0.005), but not the ASCVD risk score (p=0.588), was significantly associated with CV mortality.

### Classification of CV Mortality Across Women and Men of Diverse Race and Ethnicity

We examined separate AUC models for women and men, as reported in **Table 4**. For women, overall AUC values were high and when CAC was added to the model, there was significant improvement in classification of CV mortality (p=0.0175). Race and ethnicity did not improve the classification of CV mortality among women in a model containing the ASCVD risk score and CAC score findings (p=0.45). For men, the AUC for the ASCVD risk score was 0.78 but improved when CAC findings were added to the model (p<0.0001). In a model containing both the ASCVD risk score and CAC score findings, race and ethnicity significantly improved CV mortality classification (p=0.0021).

**Table 4.** Area Under the Curve from Receiving Operating Characteristic Models Classifying CV Mortality.

| **Model** | **Women (n=15,023)** |  | **p value** |
| --- | --- | --- | --- |
| Model #1 | ASCVD Risk Score Alone | 0.859 (0.854-0.865) | <0.001 |
| Model #2 | Adding CAC Score | 0.875 (0.869-0.880) | <0.001 |
| Model #3 | Adding Race and Ethnicity | 0.877 (0.872-0.883) | <0.001 |
|  | Model Comparison:<br>- #1 vs. #2<br>- #1 vs. #3<br>- #2 vs. #3 | p=0.0285<br>p=0.0175<br>p=0.4481 |  |
| **Model** | **Men (n=27,941)** |  | **p value** |
| Model #1 | ASCVD Risk Score Alone | 0.782 (0.777-0.786) | <0.001 |
| Model #2 | Adding CAC Score | 0.803 (0.798-0.808) | <0.001 |
| Model #3 | Adding Race and Ethnicity | 0.815 (0.810-0.819) | <0.001 |
|  | Model Comparison:<br>- #1 vs. #2<br>- #1 vs. #3<br>- #2 vs. #3 | p=0.0001<br>p<0.0001<br>p=0.0021 |  |

## Discussion

Profound racial and ethnic disparities represent significant barriers to reducing the burden of CV disease through lifesaving preventive care. Detection of atherosclerosis, through CV screening, is the basis for guiding primary prevention^12^ with CAC scanning becoming an ever important part of the screening process, especially among individuals where risk assessment is uncertain or when significant evidence gaps exist among key, diverse populations.^4,13–16^ For CAC, the evidence among racially and ethnically diverse subgroups of women and men is incomplete and often inconsistent across the published findings.^4,13–16^ With longer follow-up from a larger cohort, we identified excess CV mortality risk, especially among Black women and men as compared to other participants. The findings from this large multicenter consortium provides a few important messages: (a) There is a delayed pattern of screening leading to early and heightened CV mortality for Black women and men when compared to other subgroups. (b) Black men represent those at consistently the highest CV mortality whether having a low or higher ASCVD or CAC scores (p<0.001 for all comparisons), with Hispanic men with a 0 and ≥100 CAC score being at intermediate risk. (c) Black women and men as well as Hispanic men categorized as low risk based on the ASCVD risk or CAC scores have a higher CV mortality rate than other subgroups, suggesting residual risk that is not fully mediated by coronary atherosclerosis. (d) Finally, while clinical risk factors and CAC explain a significant proportion of the observed variability in CV mortality, the addition of race and ethnicity provides incremental risk discrimination, particularly for men. These results confirm the need for a deeper knowledge of racial and ethnic differences to drive optimal timing for CV screening and identification of unique factors aiding in CV risk detection across diverse populations of women and men.

### High CV Mortality Risk Among Black and Hispanic Women and Men

A number of series report a lower prevalence of CAC among Blacks and Hispanics as compared to other racial and ethnic counterparts; potentially categorizing these subgroups as lower risk.^17–26^ This was certainly not the case for Black women who had a ∼50% higher prevalence of CAC. A similar paradox or mischaracterization of risk has been reported among Hispanic individuals with a heavy risk factor burden yet low CV mortality (i.e., the Hispanic paradox).^27^ Earlier findings also describe a similar ability to risk stratify using CAC scores across diverse racial and ethnic subgroups but do not identify excess CV mortality risk noted herein.^1^ By comparison, we report a significantly higher overall long-term CV mortality, especially among Black women and men, with Hispanic women and men being at intermediate risk. Black women and men represent those at highest risk with a relative hazard for CV mortality of 4.6 and 3.8 (p<0.001 for both). Hispanic women and men had an intermediate risk for CV mortality ∼2-fold higher than their Asian, white, and other counterparts. These findings add uniquely given our lack of evidence as to how sex, race, and ethnicity impact the burden of CAC and ensuing CV outcomes.

For women, epidemiologic data reveal that Black women are at significantly higher risk of death due to CAD, stroke, and heart failure when compared to white women.^28^ This pattern of elevated risk is due to a greater prevalence of CV risk factors, a greater burden of atherosclerosis, and ensuing higher CV mortality^29^ has not often been reported in other CAC cohorts.^1,24,30^ We also report that Black women had the highest ASCVD risk score at the time of CAC scanning, similar to that of her male counterparts, with 40% having an ASCVD risk score >7.5%. Certainly, the screening of this high-risk subset of Black women significantly influenced their CV mortality risk, and especially so for those with a CAC score ≥100.

CV mortality was consistently highest for Black men, when compared to other men.^31,32^ In the CAC Consortium, nearly half of Black men had a high ASCVD risk score with a ∼3-fold higher relative hazard for CV death as compared to Asian, white, and other male participants. Moreover, Black men with a CAC score ≥100 had a 4-fold higher CV mortality hazard when compared to those participants identified as Asian, white, or other. We did not observe a paradox for Hispanic men who had higher CV mortality risk for those with an ASCVD risk score >7.5% or CAC score ≥100. From a single center series of more than 13,000 patients, Nakanishi and colleagues similarly reported higher 10-year all-cause mortality for Hispanic and African American patients.^33^

### Is the Power of Zero Universal?

We also uniquely reveal a higher-than-expected CV mortality among Black women and men and Hispanic men with a 0 CAC score. Much has been made of the “power of zero”^13,14,16^ as supporting a long-term warranty period for low-risk status when the CAC score is 0. Of course, the longer-term follow-up in our group would expectedly increase the number of observed CV deaths. However, similar registries have noted a low long-term mortality risk among individuals lacking CAC.^13,14,16^ Our results reveal a long-term CV mortality risk exceeding 1% for Black women and Hispanic men, but was notably highest for Black men (hazard ratio: 9, p<0.001). These results reveal the reduced life expectancy not only among Black individuals but especially so for Black men.^34^ Thus, for diverse patients with a 0 CAC score, care should be exercised when employing CV screening, especially for Black women and men and Hispanic men. Among Black women without detectable CAC, those particularly at risk included those with diabetes. High risk Black and Hispanic men with a 0 CAC score were older. These were the lone discernible high-risk characteristics among those with a 0 CAC score. Older age patients have similarly been reported to have a high risk in the setting of a 0 CAC score.^33^ Explanations for this include documentation of non-ischemic but other heart failure or vascular deaths, and a potentially greater burden of noncalcified atherosclerosis which may contribute to our findings.^35^ Importantly, our findings underscore limitations of CAC scanning when applied to broad ASCVD outcomes, especially as it relates to diverse women and men.

### Consortium Limitations

The CAC Consortium involved recruitment at 4 centers, representing unique populations undergoing CV screening, with only 3 reporting race and ethnicity. To adjust for patterns of bias and missingness, we included the enrolling site, year of scanning, and the ASCVD risk score as covariates in our prognostic models. Bias in the recording of race and ethnicity would also be reflected in the cost of the CAC scan, which was largely self-pay. Thus, although our findings add incrementally to the CAC evidence, the included subjects remain a selected subset of at-risk individuals. Importantly, baseline characteristics were similar between our consortia and the NIH-NHLBI Multi-ethnic Study of Atherosclerosis.^1^ Finally and importantly, we could not measure or quantify the impact of racism, environmental factors, or other social determinants of health to our reported racial and ethnic differences in CV mortality.

### Conclusions

With an ever-increasing diversity in our patient population, there is a growing need for greater evidence regarding effective strategies for CV screening. Both the ASCVD risk score and CAC scan are commonly used approaches to guide intensification or de-escalation of preventive therapies.^12^ Our results confirm the need for a deeper knowledge of racial and ethnic differences in CV screening especially for the optimal timing of CV screening for higher risk Black women and men contributing to worsening CV mortality. In fact, our results reveal the reduced life expectancy not only among Black women but especially so for Black men.^34^ These results support the need for expanded knowledge on not only the development of robust equations to predict CV outcomes for diverse populations but also the need to identify novel risk markers successful at stratifying diverse populations, perhaps in addition to CAC scanning.

## Data Availability

Data is available upon request and submission of specific aims.

---

**Supplemental Table 1.**
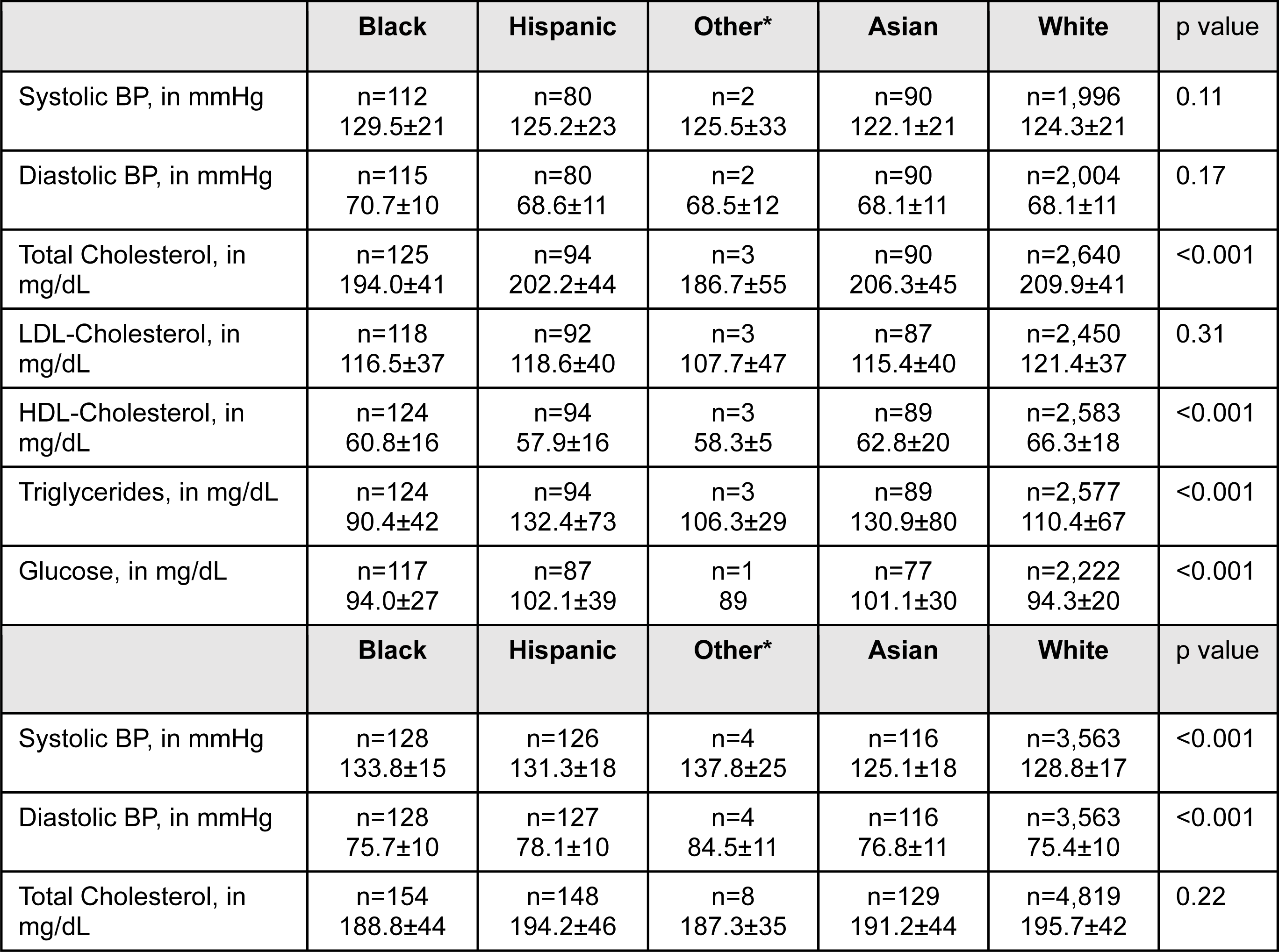
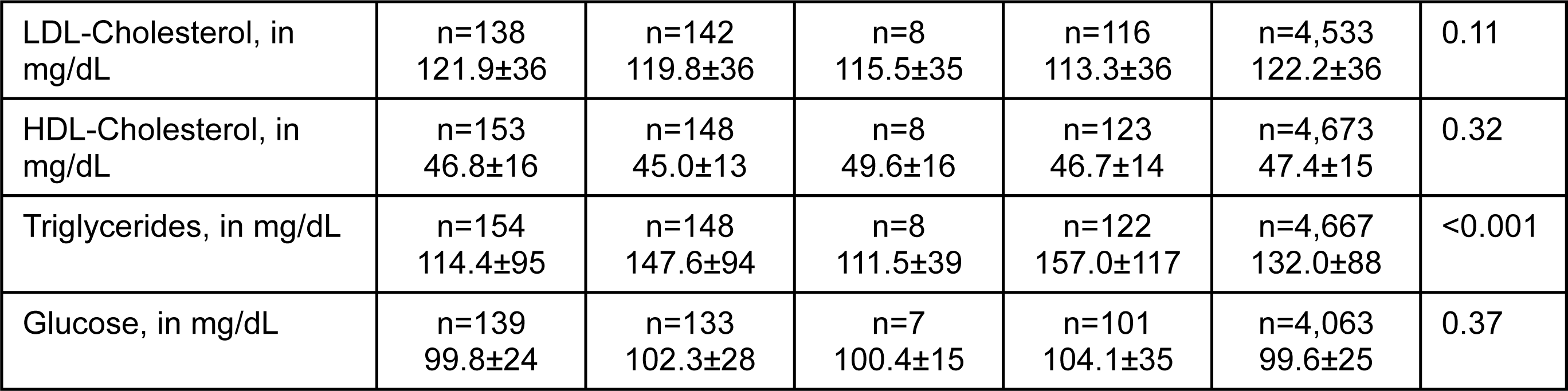
Subsample of Women and Men from the CAC Consortium with Measurements of Blood Pressure (BP), Cholesterol (Total, Low Density Lipoprotein [LDL], High Density Lipoprotein [HDL]), Triglycerides, and Glucose. Data are presented as %, except for age which is reported by median and interquartile range.

